# IL-2 and IFN-γ are biomarkers of SARS-CoV-2 specific cellular response in whole blood stimulation assays

**DOI:** 10.1101/2021.01.04.20248897

**Authors:** Begoña Pérez-Cabezas, Ricardo Ribeiro, Inês Costa, Sofia Esteves, Ana Rafaela Teixeira, Teresa Reis, Ricardo Monteiro, Alexandre Afonso, Vitor Pinheiro, Maria Isabel Antunes, Maria Lucília Araújo, João Niza Ribeiro, Anabela Cordeiro-da-Silva, Nuno Santarém, Joana Tavares

**Author notes:** Corresponding authors, Correspondence: Joana Alexandra Pinto da Costa Tavares: Instituto de Investigação e Inovação em Saúde, Rua Alfredo Allen 208, 4200-135 Porto, Portugal.; Nuno Santarém: Instituto de Investigação e Inovação em Saúde, Rua Alfredo Allen 208, 4200-135 Porto, Portugal.; Anabela Cordeiro da Silva: Instituto de Investigação e Inovação em Saúde, Rua Alfredo Allen 208, 4200-135 Porto, Portugal. Equal contribution (Inês Costa, Sofia Esteves, Ana Rafaela Teixeira, Teresa Reis).

## Abstract

A proper description of the immune response to SARS-CoV-2 will be critical for the assessment of protection elicited after both infection and vaccination. Uncoupled T and B cell responses have been described in acute and convalescent patients and exposed individuals. We assessed the potential usefulness of whole blood stimulation assays to identify functional cellular immune responses to SARS-CoV-2. Blood from COVID-19 recovered individuals (5 months after infection) and negative subjects was stimulated for 24 hours with HLA predicted peptide “megapools” of the Spike and Nucleoprotein, or the mixture of them. After stimulation, cytokines were quantified using a beads-based multiplex assay. Interleukin-2 and IFN-γ were found to be specific biomarkers of SARS-CoV-2 cellular response. Using the Spike and Nucleoprotein mixture, 91.3% of COVID-19 recovered individuals presented an IL-2 stimulation index over the cut-off, while 82.6% showed IFN-γ. All the negative individuals presented an IL-2 response under the cut-off, while 5.3% of these subjects presented positive IFN-γ stimulation indexes. Moreover, IL-2 production correlated with IgG levels for Spike 1, RBD, and Nucleocapsid. In conclusion, we demonstrate the potential of whole blood stimulation assays and the quantification of IL-2 and IFN-γ for the analysis of SARS-CoV-2 functional cellular responses.

## Introduction

Severe acute respiratory syndrome coronavirus 2 (SARS-CoV-2) is the etiological agent of the ongoing COVID-19 pandemic, responsible for more than 1.6 million deaths (WHO, 2020; Wu et al., 2020). Data from acute and convalescent patients indicated that most infected people mount antibody and T cell responses with correlated magnitudes (Braun et al., 2020; Grifoni et al., 2020; Ni et al., 2020; Peng et al., 2020; Sekine et al., 2020; Weiskopf et al., 2020). However, uncoupled T and B cell responses against SARS-CoV-2 were also described (Gallais et al., 2020; Sekine et al, 2020), which can be explained by the course of a mild infection that triggered T cell immunity without detectable antibodies, or by the waning of a transient antibody response (Altmann and Boyton, 2020; Ibarrondo et al., 2020; Long et al., 2020).

Understanding population-level immunity to SARS-CoV-2 is essential not only in the context of natural infections but also for the ongoing vaccination campaigns (WHO, 2020). Indeed, there remains a lack of clarity as to what may constitute an immunologically effective COVID-19 vaccine strategy, how to define successful endpoints in vaccine efficacy testing and what to expect from the global vaccine effort over the next few years (Jeyanathan et al., 2020). Defining biomarkers of functional cellular immune responses is particularly relevant as, similarly to human common cold coronaviruses and SARS-CoV, antibody responses to SARS-CoV-2 wane rapidly within months after infection (Ibarrondo et al, 2020; Sariol and Perlman, 2020). Therefore, control of re-infection with human coronaviruses seems mainly to be antibody independent but T cell dependent (Jeyanathan et al., 2020; Sariol and Perlman, 2020). Importantly, SARS-CoV specific T cells responses have been detected in humans 17 years after the infection (Le Bert et al., 2020). For SARS-CoV-2, specific T cells are maintained at least six months following infection being the magnitude of this response related to the clinical features (Le Bert et al., 2020; Zuo et al., 2020).

Cellular responses to SARS-CoV-2 have been mostly characterized following peripheral blood mononuclear cells (PBMCs) isolation and *in vitro* stimulation for several days with peptide pools for the quantification of IFN-γ producing cells by ELISpot or intracellular cytokine staining (Le Bert et al., 2020; Ni et al. 2020; Peng et al, 2020). To contribute to the study of functional immune responses against SARS-CoV-2, we devised a whole blood stimulation (WBS) assay to define the cytokine profile characteristic of individuals that recovered from COVID-19. The use of WBS allows the assessment of the response of circulating immune cells in a physiological environment that retains not only the whole population of leukocytes but also platelets and circulating soluble factors (Duffy et al., 2014). Moreover, whole blood is already being used in the clinics to diagnose latent tuberculosis infection through the induction of IFN-L production (Santin et al., 2012). This study contributes to the understanding of the natural immune response in mild to moderate infections which is instrumental to guide vaccine efforts and to the design of therapeutic interventions.

## Results and discussion

### Study Participants

A total of 42 healthcare workers were recruited to this study including 23 COVID-19 recovered and 19 SARS-CoV-2 negative individuals (with negative qRT-PCR in a nasopharyngeal swab performed following risk-contact tracking and monitored by IgG serology). Among the negative subjects, 9/19 (47%) were male and 10/19 (53%) were female with a median age of 37.5 years (21-60 years), while 1/23 (4.3%) COVID-19 recovered individuals were male and 22/23 (95.7%) were female with a median age of 45 years (27-61 years). Table I and supplemental Figure 1 depict COVID-19 recovered patients characteristics. COVID-19 subjects remained positive by qRT-PCR during a median of 38 days (18-106 days) and blood was collected with a median of 100 days (31-176 days) after the first negative qRT-PCR, with a total median elapsed time of 137 days (112-197 days) since the first positive qRT-PCR until the blood collection.

**Table I.**
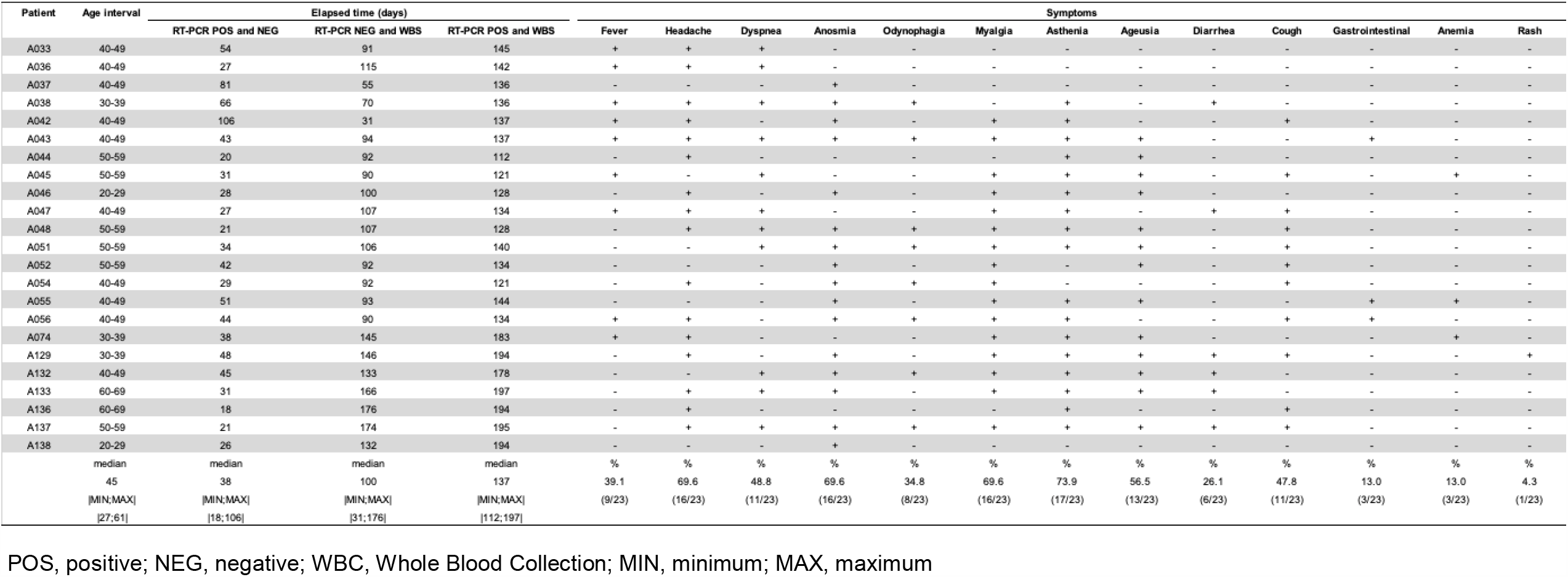
Clinical and pathological characteristics of COVID-19 recovered patients.

| Patient | Age interval | Elapsed time (days) |  |  | Symptoms |  |  |  |  |  |  |  |  |  |  |  |  |
| --- | --- | --- | --- | --- | --- | --- | --- | --- | --- | --- | --- | --- | --- | --- | --- | --- | --- |
|  |  | RT-PCR POS and NEG | RT-PCR NEG and WBS | RT-PCR POS and WBS | Fever | Headache | Dyspnea | Anosmia | Odynophagia | Myalgia | Asthenia | Ageusia | Diarrhea | Cough | Gastrointestinal | Anemia | Rash |
| A033 | 40-49 | 54 | 91 | 145 | + | + | + | - | - | - | - | - | - | - | - | - | - |
| A036 | 40-49 | 27 | 115 | 142 | + | + | + | - | - | - | - | - | - | - | - | - | - |
| A037 | 40-49 | 81 | 55 | 136 | - | - | - | + | - | - | - | - | - | - | - | - | - |
| A038 | 30-39 | 66 | 70 | 136 | + | + | + | + | + | - | + | - | + | - | - | - | - |
| A042 | 40-49 | 106 | 31 | 137 | + | + | - | + | - | + | + | - | - | + | - | - | - |
| A043 | 40-49 | 43 | 94 | 137 | + | + | + | + | + | + | + | + | - | - | + | - | - |
| A044 | 50-59 | 20 | 92 | 112 | - | + | - | - | - | - | + | + | - | - | - | - | - |
| A045 | 50-59 | 31 | 90 | 121 | + | - | + | - | - | + | + | + | - | + | - | + | - |
| A046 | 20-29 | 28 | 100 | 128 | - | + | - | + | - | + | + | + | - | - | - | - | - |
| A047 | 40-49 | 27 | 107 | 134 | + | + | + | - | - | + | + | - | + | + | - | - | - |
| A048 | 50-59 | 21 | 107 | 128 | - | + | + | + | + | + | + | + | - | + | - | - | - |
| A051 | 50-59 | 34 | 106 | 140 | - | - | + | + | + | + | + | + | - | + | - | - | - |
| A052 | 50-59 | 42 | 92 | 134 | - | - | - | + | - | + | - | + | - | + | - | - | - |
| A054 | 40-49 | 29 | 92 | 121 | - | + | - | + | + | + | - | - | - | + | - | - | - |
| A055 | 40-49 | 51 | 93 | 144 | - | - | - | + | - | + | + | + | - | - | + | + | - |
| A056 | 40-49 | 44 | 90 | 134 | + | + | - | + | + | + | + | - | - | + | + | - | - |
| A074 | 30-39 | 38 | 145 | 183 | + | + | - | - | - | + | + | + | - | - | - | + | - |
| A129 | 30-39 | 48 | 146 | 194 | - | + | - | + | - | + | + | + | + | + | - | - | + |
| A132 | 40-49 | 45 | 133 | 178 | - | - | + | + | + | + | + | + | + | - | - | - | - |
| A133 | 60-69 | 31 | 166 | 197 | - | + | + | + | - | + | + | + | + | - | - | - | - |
| A136 | 60-69 | 18 | 176 | 194 | - | + | - | - | - | - | + | - | - | + | - | - | - |
| A137 | 50-59 | 21 | 174 | 195 | - | + | + | + | + | + | + | + | + | + | - | - | - |
| A138 | 20-29 | 26 | 132 | 194 | - | - | - | + | - | - | - | - | - | - | - | - | - |
|  | median | median | median | median | % | % | % | % | % | % | % | % | % | % | % | % | % |
|  | 45 | 38 | 100 | 137 | 39.1 | 69.6 | 48.8 | 69.6 | 34.8 | 69.6 | 73.9 | 56.5 | 26.1 | 47.8 | 13.0 | 13.0 | 4.3 |
|  | [MIN;MAX] | [MIN;MAX] | [MIN;MAX] | [MIN;MAX] | (9/23) | (16/23) | (11/23) | (16/23) | (8/23) | (16/23) | (17/23) | (13/23) | (6/23) | (11/23) | (3/23) | (3/23) | (1/23) |
|  | [27;61] | [18;106] | [31;176] | [112;197] |  |  |  |  |  |  |  |  |  |  |  |  |  |

POS, positive; NEG, negative; WBC, Whole Blood Collection; MIN, minimum; MAX, maximum

### Cytokine profiling following whole blood stimulation with SARS-CoV-2 spike and nucleoprotein peptides

Effectiveness of adaptive immunity depends on the interplay of both B and T cell responses. Detection of antibody responses against SARS-CoV-2 through ELISA is well established (Zhao et al., 2019; Obka et al., 2020; To KK et al., 2020). However, a simplified and standard procedure for the identification of SARS-CoV-2 cellular responses is still not defined. Whole blood stimulation is an attractive approach due to the minimal sample handling, which is also translated into reduced variation (Duffy et al., 2014). It was successfully used to analyse the inflammatory signature to several medically relevant stimuli and is a clinical standard procedure for the identification of *Mycobacterium tuberculosis* infections (Santin et al., 2012; Duffy et al., 2014). Here, antigen-specific production of IL-5, IL-13, IL-2, IL-6, IL-9, IL-10, IFN-γ, TNF-α, IL-17A, IL-17F, IL-4, and IL-22 was measured in supernatants from WBS assays. Whole blood was stimulated during 24 hours with PepMix peptide pools for Spike (pools 1 S1, and 2-S2), or Nucleoprotein (N), or the mixture S1+S2+N and the cytokines were quantified in the plasma using a beads-based assay (supplemental Table I). The Spike (S) protein of SARS-CoV-2 is separated at the cleavage site into subunit S1, which comprises the receptor binding domain (RBD) and subunit S2 which contains the phusion peptide, the transmembrane domain and the cytoplasmic peptide. The S1 pool spans the N-terminal amino acid (aa) residues 1-643, while the S2 pool covered the C-terminal aa residues 633-1273. The N pool covered 419 aa of the Nucleoprotein (Braun et al., 2020). No differences in the cytokines production in response to stimuli such as Phytohemagglutinin-A (PHA) and Staphylococcus enterotoxin B (SEB) were seen between COVID-19 recovered and negative subjects (supplemental Table I; data not shown). A significant increase upon SARS-CoV-2 peptide pools stimulation was only seen for IL-2, IFN-γ, IL-6, and TNF-α (Figure 1). IL-2 and IFN-γ were significantly produced by COVID-19 recovered individuals in response to all peptide pools when compared to unstimulated condition. Negative subjects also showed a slight but significant increase in the production of both IL-2 and IFN-γ only after stimulation with the mixture S1+S2+N. Despite in lower amounts, the production of IFN-γ is also significant in negative individuals when stimulated with S1. An increased production upon stimulation with the S1+S2+N mixture is also seen for IL-6 and TNF-α in both, the negative and the COVID-19 recovered groups (Figure 1). Stimulation with the S2 peptide pool leads to a significant increase in the production of IL-6 in COVID-19 recovered subjects, while stimulation with S2 and N peptide pools induces significant production of TNF-α (Figure 1).

**Figure 1.**
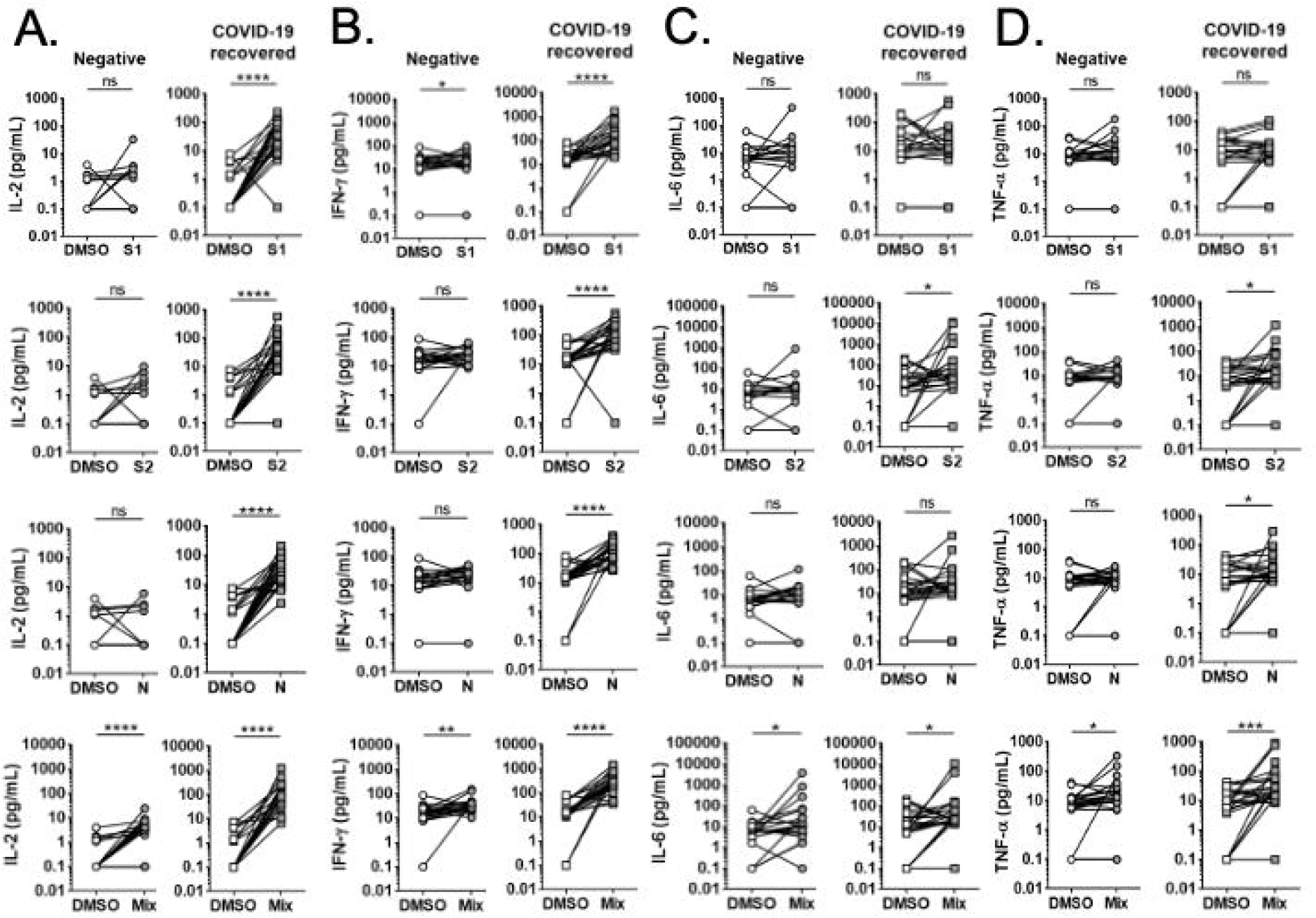
IL-2, IFN-γ, IL-6 and TNF-α quantification after WBS assays from negative and COVID-19 recovered patients, in response to SARS-CoV-2 PepMix peptide pools. Graphs representing the production of IL-2 (A), IFN-γ (B), IL-6 (C) and TNF-α (D) measured after WBS with SARS-CoV-2 peptide pools. Whole blood from negative (n=19), COVID-19 recovered individuals (n=23), and asymptomatic patients (n=3) was stimulated in vitro for 24 h with peptide pools of SARS-CoV-2 (1 µg/mL per peptide) Spike S1 (S1), Spike S2 (S2), Nucleocapsid (N) or a mixture of S1+S2+N. After that, the supernatant was collected and frozen. Cytokine concentration was determined by flow cytometry using a LEGENDplex Multiplex kit. Each symbol pair connected by a line represents a different individual. White symbols indicate the cytokine concentration produced in response to the negative control situation (DMSO), while grey symbols represent the cytokine concentration produced in response to the indicated peptide pool. Statistical differences were inferred using the Wilcoxon signed-rank test; ns-non-significant; p≤0.05*; p≤0.01**; p≤0.001***; p≤0.0001****.

To quantify the intensity of the response, stimulation indexes (SI) were calculated by dividing the cytokine concentration produced by each individual in response to the peptide pools, by the cytokine concentration secreted by the same individual in response to the control condition (DMSO). When comparing the stimulation indexes obtained in the COVID-19 recovered patients with the ones from the negative group, IL-2 and IFN-γ were the only cytokines significantly produced by COVID-19 recovered subjects in response to the peptide pools (Figure 2). This production was consistent for almost all the COVID-19 recovered individuals (Figure 2). Combination of S1+S2+N was the stimuli that generated a significantly higher stimulation index (SI) for both IL-2 and IFN-γ (supplemental Figure 2). Stimulation indexes for IL-2 and IFN-γ obtained using S1+S2+N significantly and positively correlate (p=0.0389; r=0.4334) (supplemental Figure 3A). Considering the significant difference between the negative and COVID-19 subjects receiving operating curves (ROC) were used to define cut-offs. The areas under the curve for IL-2 were consistently higher than the ones for IFN-γ (supplemental Table II). As expected, the overall performance of S1+S2+N was as high or better than any individual stimuli (supplemental Table II). Thus, specificity and sensitivity for the IL-2 response in this condition was 91.3% (21/23) and 100% (19/19), respectively, while for IFN-γ were 82.6% (19/23) and 94.7% (18/19). Regarding the individual stimuli and IL-2 response, 100% specificity and 91.3% sensitivity is also achieved with the S2 and N peptide pools, while for IFN-γ the higher performance was obtained with N stimulation (89.5% specificity; 82.6% sensitivity) (supplemental Table II). Population cut-offs, calculated using the mean ± 2 standard deviations (2SD) of the SI obtained from the negative group, for each stimuli and cytokine, did not improve the performance of the assay (data not shown). The specific production of IL-2 and IFN-γ to SARS-CoV-2 Spike and Nucleoprotein is in agreement with other’s findings (Peng et al., 2020).

**Figure 2.**
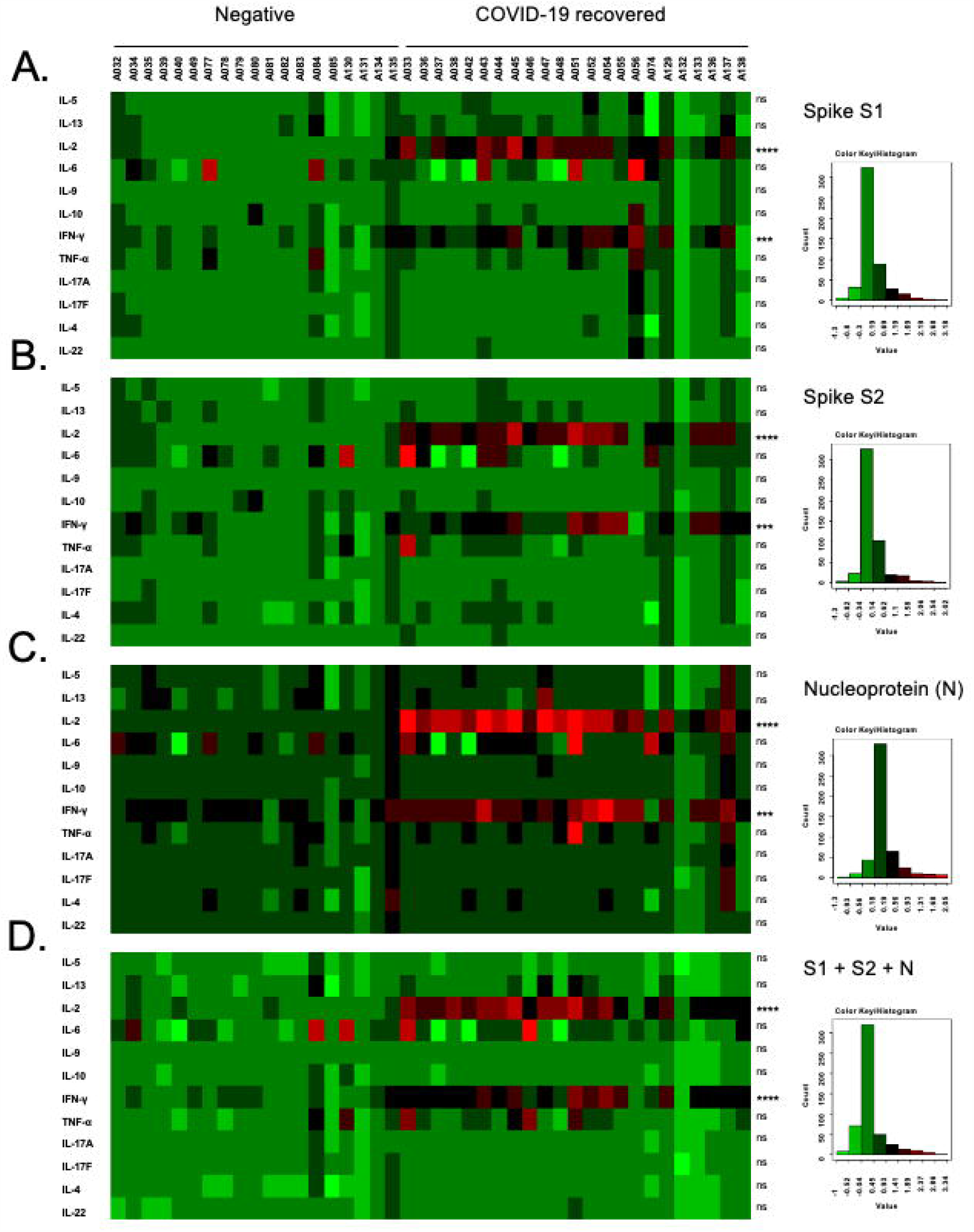
Cytokines stimulation indexes in whole blood stimulation assay for SARS-CoV-2. Heatmaps representing the cytokine response measured after whole blood stimulation with SARS-CoV-2 PepMix peptide pools. Whole blood from negative (n=19) and COVID-19 recovered individuals (n=23) was stimulated *in vitro* for 24 h with PepMix peptide pools (1 μg/mL per peptide) of SARS-CoV-2 Spike (S1 and S2), Nucleocapsid (N) or a mixture of S1+S2+N. After that, plasma was collected and frozen. Cytokine concentration was determined by flow cytometry using a LEGENDplex Multiplex kit. Each column represents a different individual. Each row indicates the stimulation index (SI) for each cytokine and individual (Log-10 scale). The stimulation index was calculated dividing the cytokine concentration obtained after each stimulation by the quantification obtained in the negative control (DMSO). Statistical differences were inferred using a Kruskal-Wallis test; ns-non-significant; p≤0.001***; p≤0.0001****.

Interestingly, in the conditions used, IL-2 performed better than IFN-γ, showing higher sensitivity and specificity for any of the antigens used. Interleukin-2 is a growth factor for cellular expansion of specific T cells and the generation of effector and memory T cells (Abbas et al., 2018). It has been previously detected upon *in vitro* stimulation of PBMCs from both acute and convalescent COVID-19 patients (Braun et al., 2020; Grifoni 2020 et al., 2020; Peng et al., 2020; Sekine et al., 2020; Thieme et al., 2020; Weiskpof et al., 2020). Intracellular staining revealed that both CD8+T and CD4+ T cells are sources of this cytokine but CD4+T cells appear to be the main producers (Braun et al., 2020; Peng et al., 2020; Sekine et al., 2020; Thieme et al., 2020). However, IL-2 quantification after WBS was never proposed for the identification and stratification of SARS-CoV-2 cellular responses.

Importantly, IL-2 SI negatively correlated (r=-0.4234, p= 0.0441) with the elapsed time between the first positive qRT-PCR and the moment of the WBS assay (supplemental Figure 3B), suggesting that the detection of cellular responses against SARS-CoV-2 could be influenced by time. SARS-CoV-specific T cells have been detected in the periphery 17 years after the infection upon a previous *in vitro* expansion (Le Bert et al., 2020). The same might be also required for the study of SARS-CoV-2 cellular response in samples collected after long periods of time post-infection.

In conclusion, a specific and robust IL-2 and IFN-γ response is seen 5 months after infection, in COVID-19 recovered individuals. Whether such a high performance for the IL-2 and IFN-γ responses will be observed in populations from different geographical areas, where different circulating coronaviruses (such common cold coronaviruses, but also other unknown coronaviruses of animal origin) have been previously present remains to be addressed (Le Bert et al., 2020). Indeed, emerging evidences indicate that CD4+T cells in 35% of healthy individuals not exposed to SARS-CoV-2 recognized the Spike protein and that 40-60% of CD4+T cells in unexposed individuals are reactive to SARS-CoV-2 proteins other than Spike (Grifoni et al., 2020; Braun et al., 2020).

### IL-2 and IFN-γ mediated cellular response correlates with SARS-CoV-2 specific antibodies

To assess relationships between specific IL-2 and IFN-γ production in WBS with the humoral response, an in-house IgG ELISA was optimized for exposure detection using S1, RBD and N as antigens (Supplemental Figure 4). Seropositivity of the COVID-19 recovered cohort was found to be 95.6% for S1, 82.6% for RBD and 73.9% for N (Figure 3A). Importantly, significant correlations between IL-2 response to S1+S2+N stimuli and IgG to S1 (p=0.0004; r=0.5185), RBD (p=0.0004; r=0.5185) and N (p=0.0004; r=0.5185) were observed, unlike what was observed for IFN-γ (Figure 3B). When looking at the individual stimuli, and in agreement with existing IFN-γ ELISpot data (Peng et al. 2020; Zuo et al. 2020), significant correlations between the IFN-γ response following WBS with S1 peptides, and IgG to S1 (p=0.0058; r=0.5566) and RBD (p=0.0022; r=0.6065) were seen (Supplemental Figure 5).

**Figure 3.**
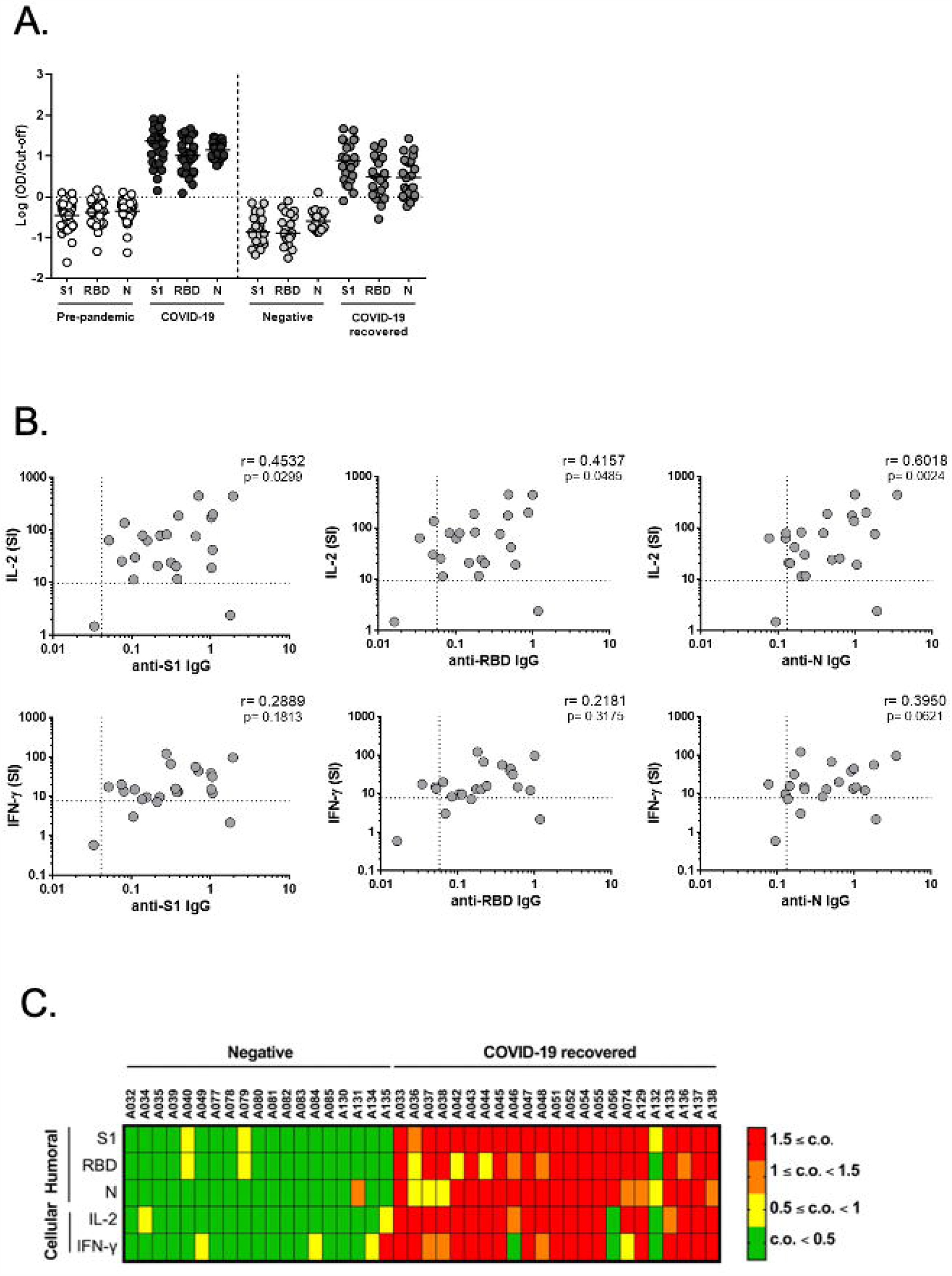
Combination of IL-2 and IFN-γ production in whole blood stimulation assays with anti-SARS-CoV-2 serology. (A) Seropositivity to SARS-CoV-2 antigens: Spike subunit S1 (S1), Receptor Binding Domain (RBD) and Nucleocapside (N) in four distinct cohorts: pre-pandemic (n=43); COVID-19 (n=30); COVID-19 recovered (n=23) and negative (n=19). In each column, a dot represents the logarithmized cut-off normalized average of at least two independent ELISA determinations, performed in duplicate, for a unique individual. The horizontal dashed line represents the cut-off. (B) Correlations of IL-2 and IFN-γ stimulation index (SI) in whole blood stimulation assays from COVID-19 recovered individuals (n=23) in response to PepMix S1+S2+N peptide pools mixture, and SARS-CoV-2 specific IgG (OD values) for S1, RBD and N. Dashed lines represent the cut-off values (IL-2 and IFN-γ calculated by ROC analysis; ELISA IgG 2SD obtained from the pre-pandemic population). The r values were calculated using Pearson’s Correlation Test. (C) Summary map for the response intensity to each studied parameter. Top panel, red label represents seropositivity or IL-2 and IFN-γ SI ≥ 1.5 cut-offs; orange label represents seropositivity or IL-2 and IFN-γ SI above the cut-off and < 1.5 cut-offs; yellow label represents seronegative samples or IL-2 and IFN-γ SI that are under the cut-offs and ≤ 0.5 cut-offs; green label represents seronegative samples or IL-2 and IFN-γ < SI 0.5 cut-offs.

The comparison of the humoral and cellular responses among COVID-19 recovered and negative subjects is represented in Figure 3C. The label represents the following seropositivity or IL-2 and IFN-γ SI: red (above 1.5 cut-off), orange (bellow 1.5 and above 1 cut-offs), yellow (bellow 1 and higher than 0.5 cut-off) and green (bellow 0.5 cut-off). Six COVID-19 recovered individuals (A36, A37, A38, A42, A44, A132) were not seropositive to all of the three antigens. Concerning the cellular response, IL-2 SI was below the cut-off for only two individuals (A56, A132), while for IFN-γ, four subjects presented a SI under the cut-off (A46, A56, A74, A132). Of note, A56 subject presented serological response without detecatble cellular response and A132 shows no indicators for either humoral or cellular response (Figure 3C). This is in agreement with previously reported data (Sekine et al., 2020; Gallais et al., 2020). However, we cannot exclude that these individuals have SARS-CoV-2 specific cells targeting other than Spike and Nucleoprotein antigens or that, alternatively, virus specific cells to Spike and Nucleoprotein do not produce or are low-producers of IL-2 and IFN-γ.

In summary, stimulation of whole blood from COVID-19 recovered individuals with SARS-CoV-2 peptide pools led to consistent production of IL-2 and IFN-□. However, IL-2 SI presented better discriminatory potential between negatives and COVID-19 recovered individuals than IFN-γ, reflected by higher sensitivity and specificity percentages with any of the peptide pools used. Despite the limited number of individuals studied, these observations highlight the potential of the whole blood assay for the identification of SARS-CoV-2 cellular responses through the production of IFN-γ and IL-2. Whether or not these specific cellular responses play a role in immune protection merits further investigation in larger prospective cohorts as there are currently no immune correlates of protection for SARS-CoV-2 or other human coronaviruses (Jeyanathan et al., 2020). Whereas the current successful vaccines to influenza and measles depend largely on antibody responses, emerging evidences suggests the requirement of both antibody and T-cell mediated immunity for SARS-CoV-2 protection (Tay et al., 2016; Sariol and Perlman, 2020). Most vaccines are designed to induce antibodies to the SARS-CoV-2 spike protein, but it is not yet known if this will be sufficient to induce full protective immunity to SARS-CoV-2 (Folegatti et al., 2020; Krammer, 2020; Zhu et al., 2020; Vorsey et al., 2020). Studying natural immune response to the virus, including the specific cellular responses, is thus critical to fill the current knowledge gaps for improved vaccines design (Jeyanathan et al., 2020).

In conclusion, this study highlights the potential use of IL-2 and IFN-γ quantification following WBS to easily define the presence of specific cellular responses not only upon natural infection but also after vaccination

## Materials and methods

### Study Design

Here, we set out to characterize the cytokines profile using a whole blood stimulation (WBS) assay combined with virus specific IgG (to SARS-CoV-2 Spike S1 subunit (S), Receptor Binding Domain (RBD) and Nucleocapsid (N)) in samples from COVID-19 recovered subjects (n=23). The median time from the negative qRT-PCR on a nasopharyngeal swab to blood collection was 100 days. Negative SARS-CoV-2 group (n=19) included subjects with a negative qRT-PCR performed following risk-contact tracking and monitored by IgG serology (Alinity i, Abbott). This study relied on the use of pre-designed mega pools containing overlapping peptides or predicted epitopes for T cell stimulation of SARS-CoV-2 spike (S1 and S2 pools) and Nucleoprotein (9). Serum samples from COVID-19 patients three weeks after symptoms onset (n=30) and a pre-pandemic (n=43) cohort were used for in-house ELISA cut-off’s determination.

### Study Approval

This study was approved by the Ethics Committees of Centro Hospitalar e Universitário de Coimbra (CHUC-084-20). Participants signed an informed consent form, according to the Helsinki principles, the Oviedo Convention and according to the General Data Protection Regulation – Regulation (EU) 2016/679.

### Study Participants

Individuals from the Coimbra region of Portugal, followed up in the Occupational Health Department at the Centro Hospitalar e Universitário de Coimbra (CHUC), diagnosed with COVID-19 (confirmed by positive qRT-PCR on a sample from the respiratory tract) and recovered (confirmed by at least two negative qRT-PCR on a nasopharyngeal swab), and a group of negative individuals (tested negative to qRT-PCR for SARS-CoV-2, performed following risk-contact tracking, and monitored routinely by serology to SARS-CoV-2 at the Occupational Health Department-CHUC) were invited to participate in this study. Blood was collected by venipuncture of the forearm, following routine clinical follow-up visits (negative and COVID-19 recovered). Among the negative subjects, the median cohort age was 37.5 years (21-60 years), 9/19 (47%) were male, and 10/19 (53%) were female. Among the COVID-19 recovered individuals, the median cohort age was 45 years (27-61 years), 1/23 (4.3%) were male, and 22/23 (95.7%) were female.

Cut-offs for the in-house ELISA were determined using SARS-CoV-2 seropositive (in Snibe; Euroimmun and Abbott tests) samples collected three weeks after COVID-19 symptoms and pre-pandemic samples. The SARS-CoV-2 samples were obtained from patients living in the Coimbra region selected from banked serum at the Clinical Pathology Department of CHUC, as recommended by the Portuguese General Health Department (Direção Geral de Saúde). Banked pre-pandemic (2000/2001) serum/plasma samples from healthy Portuguese individuals stored at 80°C were obtained from the Pharmacy Faculty of Porto University.

### Data Reporting

No statistical methods were used to predetermine sample size. The investigators were blinded to sample allocation during experiments.

### Blood collection and processing

Venous blood was collected into lithium-heparin tubes (Becton Dickinson, CA, USA). For the WBS assay, blood was used within 3 hours collection. Blood was centrifuged at 2000 g for 10 min at 20 °C, and plasma was collected, aliquoted and frozen at −80 °C until tested for ELISA determinations.

### SARS-CoV-2 Peptide pools

The PepMix™ SARS-CoV-2 Spike Glycoprotein (UniProt: P0DTC2) pool (S1), PepMix™ SARS-CoV-2 Spike Glycoprotein pool (S2), and PepMix™ SARS-CoV-2 NCAP (UniProt: P0DTC9; N) with at least 70% purity by LC-MS for each peptide (JPT Peptide Technologies Inc., MA, USA) were used in WBS assays. The PepMix ™ SARS-CoV-2 spike glycoprotein S1 pool covered the N-terminal amino acid (aa) residues 1-643 and contained 158 15-mer peptides overlapping by 11 aa, while the S2 pool covered the C-terminal aa residues 633-1273 containing 156 15-mer peptides, overlapping by 11 aa, and one 17-mer peptide at the C-terminus (157 peptides in total). The PepMix™ SARS-CoV-2 NCAP covered 419 aa of the Nucleoprotein, containing 102 15-mer peptides with 11 aa overlap. All peptide pools were reconstituted in 50 μl DMSO according to the manufacturer instructions (0.5 mg/mL) and used at 1 ug/mL per peptide as previously reported (Braun et al., 2020).

### Whole blood stimulation assay

Within 3 h of collection, 500 μl of blood was transferred to 2 mL eppendorfs. The DMSO concentration was adjusted to 0.6% in all the tested peptide pools conditions and positive controls. The assay consisted of three tubes containing 1 μL of the peptide pool (S1, S2 or N), one tube containing a mixture of 1 μL of each peptide pool (S1+S2+N), positive controls, containing either 0.5 μg of SEB (Toxin Technology, Inc., FL, USA) or 5 μg of PHA (Sigma-Aldrich, MO, USA), negative controls with DMSO. All tubes were incubated at 37 °C for 24 h. Then, supernatants (plasma) were collected and stored at −80 °C until cytokine quantification.

### Cytokine determination

Cytokines IL-5, IL-13, IL-2, IL-6, IL-9, IL-10, IFN-γ, TNF-α, IL-17A, IL-17F, IL-4, and IL-22 were quantified in supernatants from WBS assays using the Human T Helper Cytokine Panel Version 2 (12-plex) LEGENDplex kit (BioLegend, CA, USA), as recommended by the manufacturer. Samples were acquired in a LSRFortessa™ flow cytometer (BD Biosciences CA, USA) using the FACSDiva™ software v8.0 (BD Biosciences). Analysis was performed using the LEGENDplex analysis software v8.0 (BioLegend). Data were obtained in pg/mL. Stimulation indexes (SI) were calculated by dividing the cytokine concentration produced by each individual in response to the peptide pools, by the cytokine concentration produced by the same individual in response to the negative control condition (DMSO).

### SARS-CoV-2 recombinant proteins

The protein antigens used for ELISA were purchased from Sino Biological (PA, USA). Nucleocapsid-His recombinant Protein (YP_009724397.2(335Gly/Ala)) (Met1-Ala419) expressed in Baculovirus-Insect Cells with a polyhistidine tag at the C-terminus (Reference 40588-V08B); Spike protein S1 Subunit (YP_009724390.1) (Val16-Arg685) expressed in HEK293 Cells with a polyhistidine tag at the C-terminus (Reference 40591-V08H); Spike protein Receptor Binding Domain (RBD) (YP_009724390.1) (Arg319-Phe541) expressed in HEK293 Cells with a polyhistidine tag at the C-terminus.

### Serology

Ninety-six-well flat-bottom microtiter plates (Greiner® Bio-One, Austria) were coated with 50 μL of 0.05 M carbonate buffer, pH=9.6, containing 2.5 μg/mL of either spike surface glycoprotein S1 subunit (S1), Receptor Binding Domain (RBD) or Nucleocapsid protein (N). Each plate contained the three individual antigens occupying parallel duplicate lines in the centre of the plate (22 wells/antigen). Plates were incubated overnight at 4°C and blocked with 200 μL of PBS low-fat milk (10%) at 37°C for at least 1 h. Then, plates were washed in a Microplate Washer (Thermo Scientific) with PBS-Tween 20 (Sigma-Aldrich) (PBS-T 0.05%). Sera samples were diluted at 1:100 in PBS-T low-fat milk (1%) and dispensed using a single dilution for the three antigens. As an internal positive control for nucleocapsid, a pool of sera from several seropositive human samples was diluted at 1:1000 in PBS-T low-fat milk (1%). As a positive control for S1 and RBD, it was used a chimeric monoclonal antibody (Sino Biological) diluted at 1:10000 in PBS-T low-fat milk (1%). Plates were incubated with the samples at 37°C for 1 h. After a washing step, 100 μL anti-human IgG antibody conjugated with horseradish peroxidase (Sigma-Aldrich) diluted at 1:5000 was added and the plates were incubated at 37 °C for 1 h. Plates were washed and then incubated at room temperature for 10 min in dark with 0.5 mg/mL of o-phenylenediamine dihydrochloride (Sigma-Aldrich) in 0.05 M of citrate buffer, pH=5.3. As a reactive oxygen metabolic byproduct, 10 µL of hydrogen peroxide 30% (Merck, Germany) were used. Reaction was stopped with 50 μL of hydrochloric acid (3 M) (Fisher Chemical, MA, USA). Absorbance was read at 492 nm in the automatic reader Synergy2 (BioTek Instruments Inc., VT, USA). All samples were assayed as duplicate in at least two independent assays.

### Data and Statistical analysis

Microsoft Excel v.14.1.0 (Microsoft Corporation, WA, USA) and Prism 7 (GraphPad software, CA, USA) were used for plotting and statistical analysis. Statistical and significance details are provided in the respective figure legends. Individual SI heat maps were created using the heat map tool of the HIV database http://www.hiv.lanl.gov/.

Differences in cytokine SI among negative and COVID-19 recovered individuals were inferred using Kruskal-Wallis test, while differences among unstimulated and stimulated samples from COVID-19 recovered individuals were analysed with Wilcoxon signed-rank test for paired comparisons. Correlation analyses were performed using Pearson’s Correlation Test.

Receiver operating characteristic (ROC) curves for IL-2 and IFN-γ were generated following their quantification in the supernatants of WBS assay from COVID-19 recovered and negative cohorts. A 95 % confidence interval (95 % CI) for the area under the ROC curve was considered.

## Data Availability

All data referred to in the manuscript is available upon request to corresponding authors.

## Author Contributions

JT, NS and ACS conceived the study; JT, NS and BPC designed the experiments; BPC, IC, SE, ART, TR, RM and NS performed experiments; RR, AA, VP, TR, MLA and MIA were involved in patient recruitment, sample collection and clinical reporting; BPC, RR, NS, JT, ACS and JNR analysed the data and discussed the results; BPC, JT, NS and ACS wrote the manuscript and all the authors contributed to its editing.

## Acknowledgements

This work was financed by the Portuguese Foundation for Science and Technology (FCT) in the framework of the project RESEARCH4COVID 19 reference number 460. This work also received funds through the Research Unit 4293. Individual funding from FCT through CEECIND/02362/2017 (to JT). NS and BPC are funded respectively through the FCT projects PTDC/SAU-PAR/31013/2017 and PTDC/SAU-PAR/31340/2017. FCT funded PhD scholarships SFRH/BD/140177/2018 (to IC) SFRH/BD/140119/2018 (to SE) and SFRH/BD/133276/2017 (AT).

## Abbreviations

CHUC: Centro Hospitalar Universitário de Coimbra
N: Nucleoprotein
PBMC: Peripheral Blood Mononuclear Cells
ROC: Receiving Operating Curves
S: SARS-CoV-2 Spike S1 subunit
SEB: Staphylococcus enterotoxin B
SI: Stimulation Index
S1: Spike PepMix peptide pool S1
S2: Spike PepMix peptide pool S2
WBS: Whole Blood Stimulation

## Supplemental data

### Supplemental Figures

**Supplemental Figure 1.**
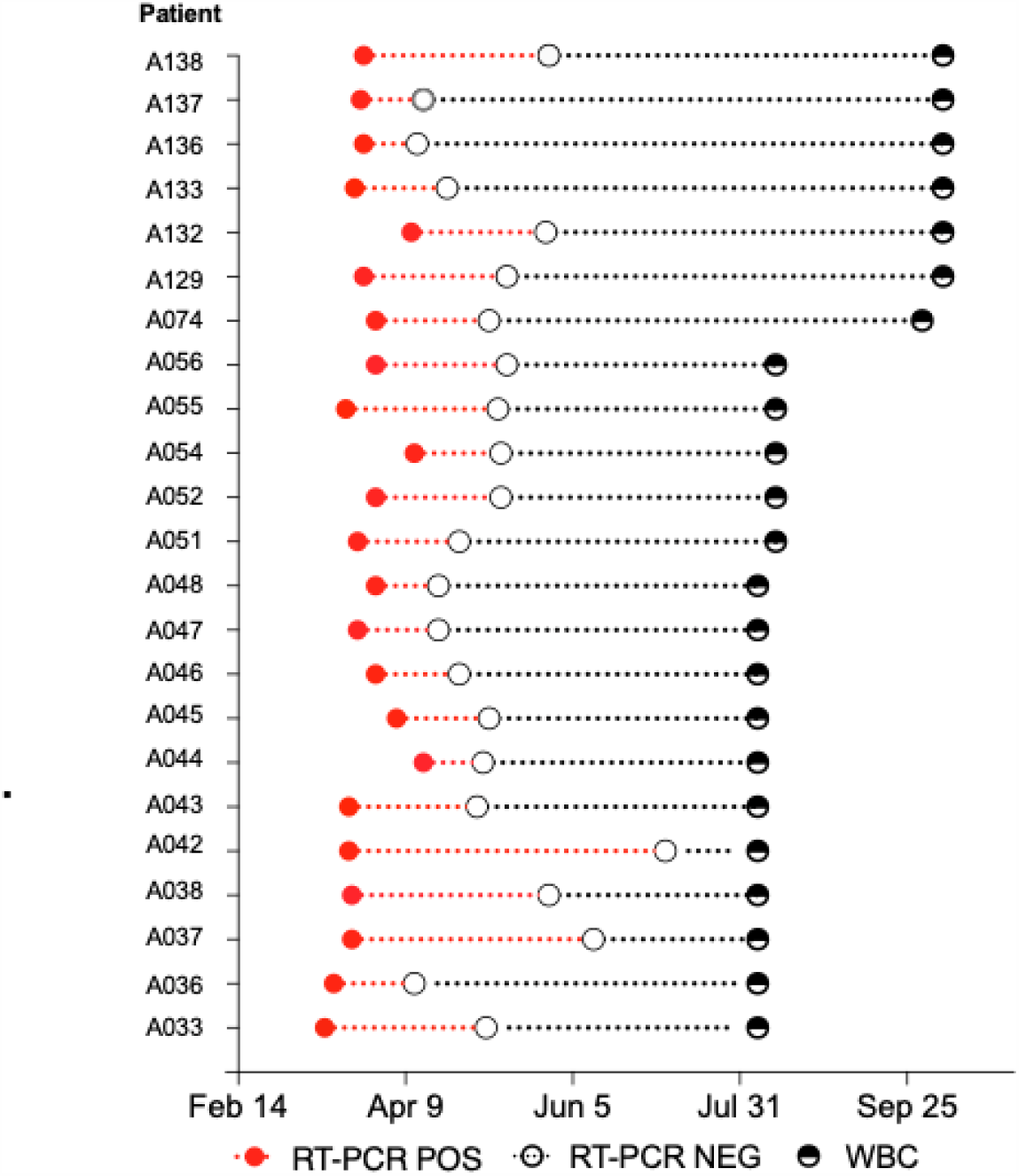
Schematic representation of the elapsed time of the COVID-19 recovered patients (2020). WBC: whole blood collection; RT-PCR POS: RT-PCR positive; RT-PCR NEG: RT-PCR negative.

**Supplemental Figure 2.**
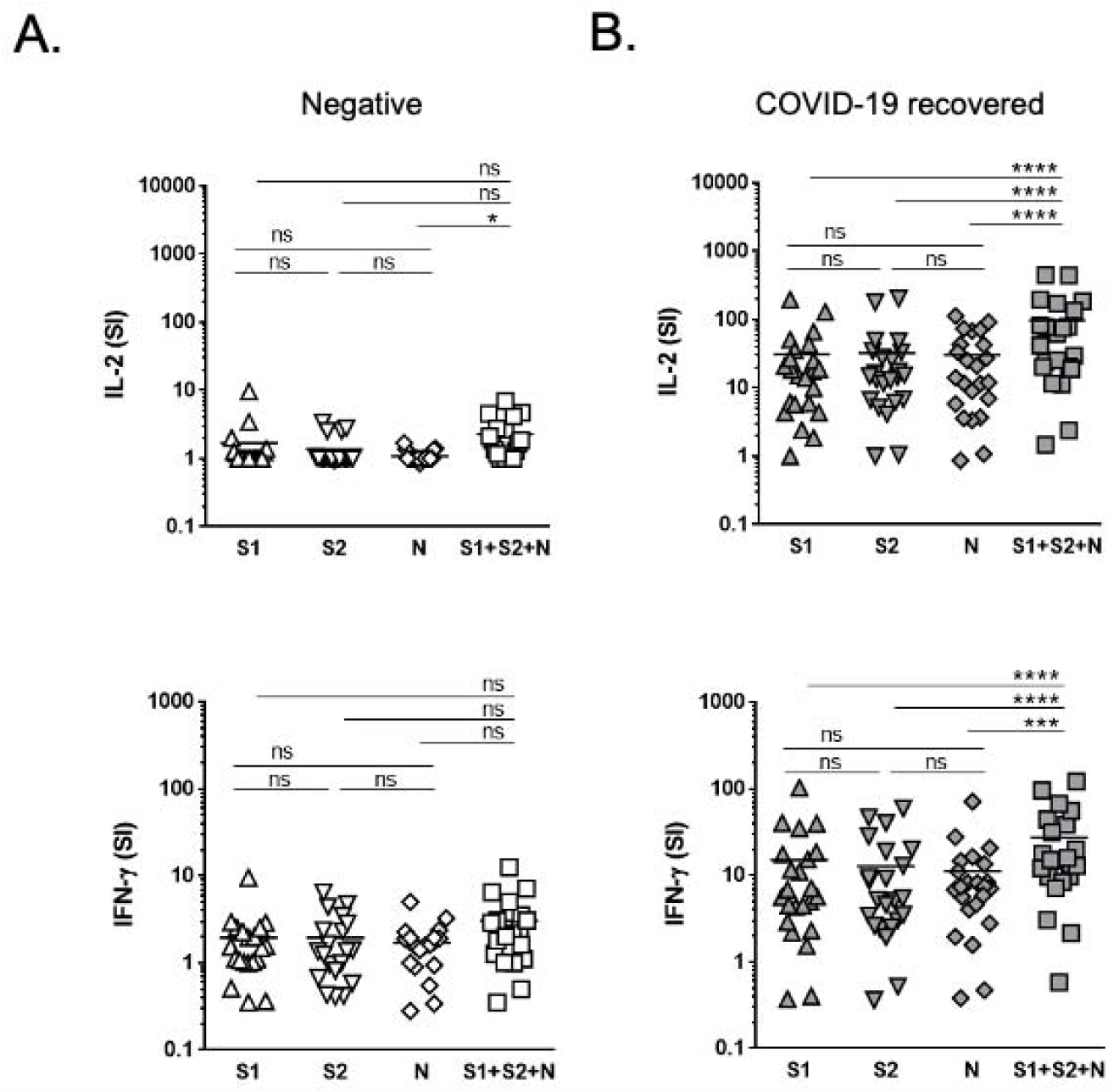
IL-2 and IFN-γ SI for SARS-CoV-2 Spike and Nucleoprotein whole blood stimulation assay. Whole blood from negative (n=19) and COVID-19 recovered individuals (n=23) was stimulated *in vitro* for 24 h with PepMix peptide pools (1 μg/mL per peptide) of SARS-CoV-2 Spike pool S1, Spike – pool S2, Nucleoprotein (N) or a mixture of S1+S2+N. After that, plasma was collected and frozen. Cytokine concentration was determined by flow cytometry using a LEGENDplex Multiplex kit. Stimulation indexes for IL-2 and IFN-γ were calculated dividing the cytokine concentration obtained after each peptide pool stimulation by the quantification obtained in the negative control (DMSO). Each symbol represents a different individual. The horizontal line in each column indicates the mean of the SI for each stimulation (Log-10 scale). Statistical differences were inferred using a Friedman test; ns-non-significant; p≤0.05*; p≤0.001***; p≤0.0001****.

**Supplemental Figure 3.**
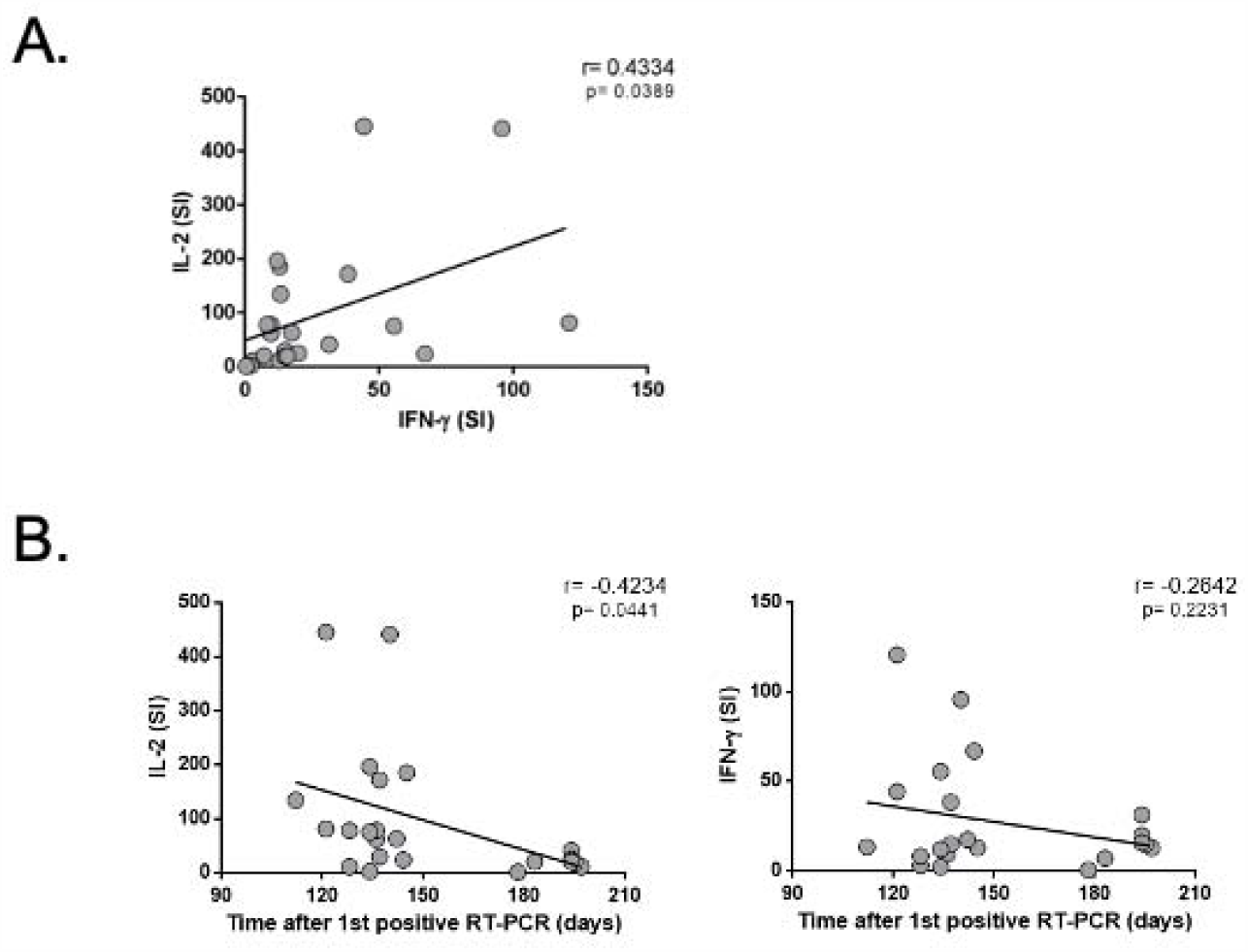
Correlations between IL-2 and IFN-γ production with the time after SARS-CoV-2 infection. (A) Linear regression of IL-2 and IFN-γ production represented by the stimulation indexes obtained after WBS assays from COVID-19 recovered individuals (n=23) in response to S1+S2+N peptide pools mixture. r values were calculated using Pearson’s Correlation Test. (B) Linear regressions of IL-2 or IFN-γ production stimulation indexes in WBS assays from COVID-19 recovered individuals (n=23) in response to S1+S2+N peptide pools mixture, and the time passed between the first positive qRT-PCR for the presence of SARS-CoV-2 and the whole blood collection for the performance of WBS assays. r values were calculated using Pearson’s Correlation Test.

**Supplemental Figure 4.**
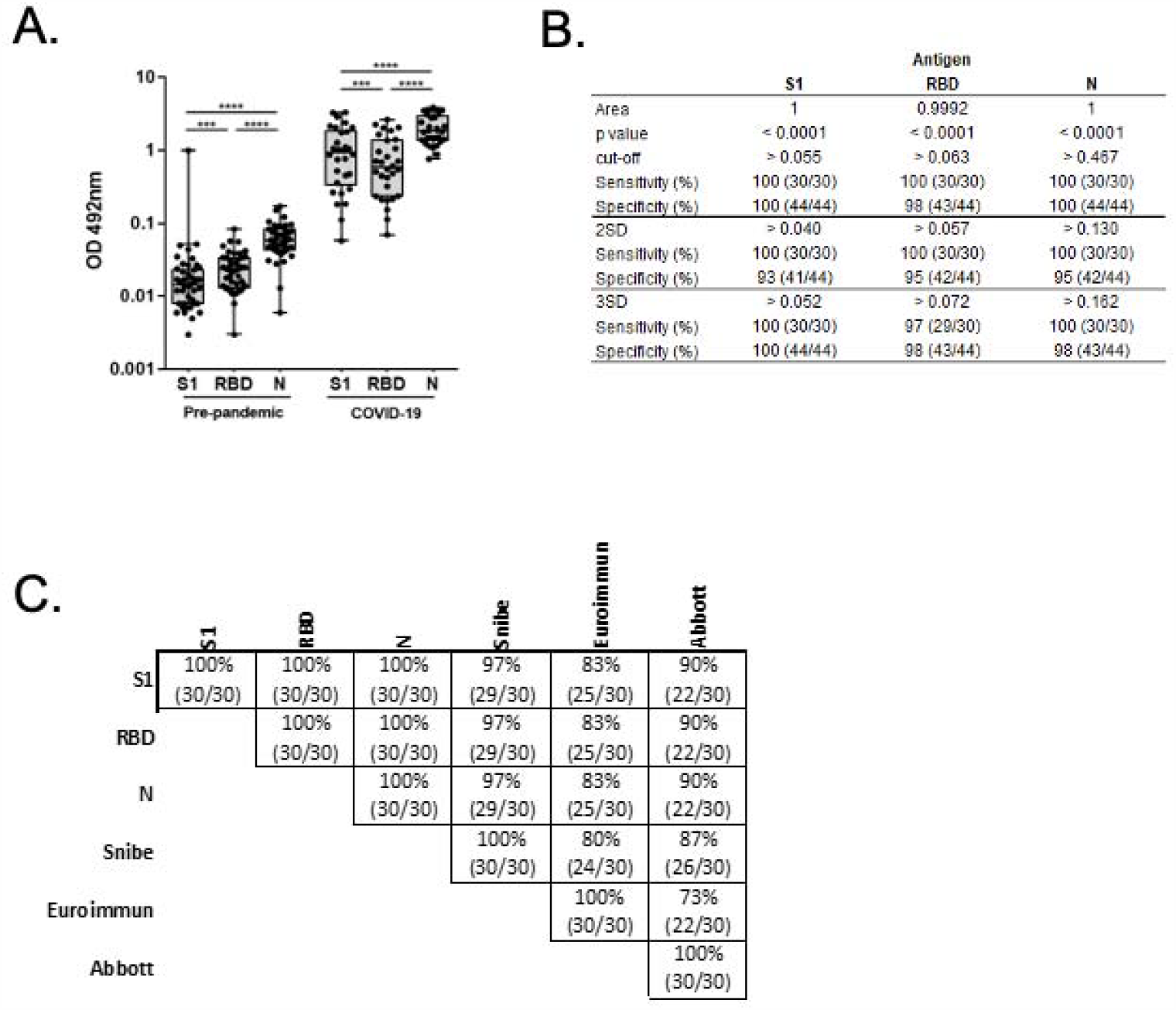
In-house SARS-CoV2 ELISA. (A) Maximum, minimum and interquartile range for specific sera reactivity, as determined by the OD 492nm in IgG ELISA assay, for SARS-CoV-2 antigens: spike subunit S1 (S1), Receptor binding domain (RBD) and Nucleoprotein (N) in pre-pandemic (n=43) and in COVID-19 (n=30) samples. In each column, a dot represents the average of at least two independent ELISA determinations, performed in duplicate, for a unique individual. Inter-cohort antigen-specific variability was evaluated using Tukey’s multiple comparison test. p≤ 0.001***; p≤ 0.0001****. (B) ROC curves, cut-offs, sensitivity, and specificity for the in-house IgG ELISA for S1, RBD, and N antigens were determined in the pre-pandemic and COVID-19 cohorts. (C) Percentage of agreement between the in-house ELISA for IgG to S1, RBD, and N and three commercial serological tests (Snibe Spike and Nucleoprotein, Euroimmun – Spike S1, Abbot Nucleoprotein) in the COVID-19 cohort.

**Supplemental Figure 5.**
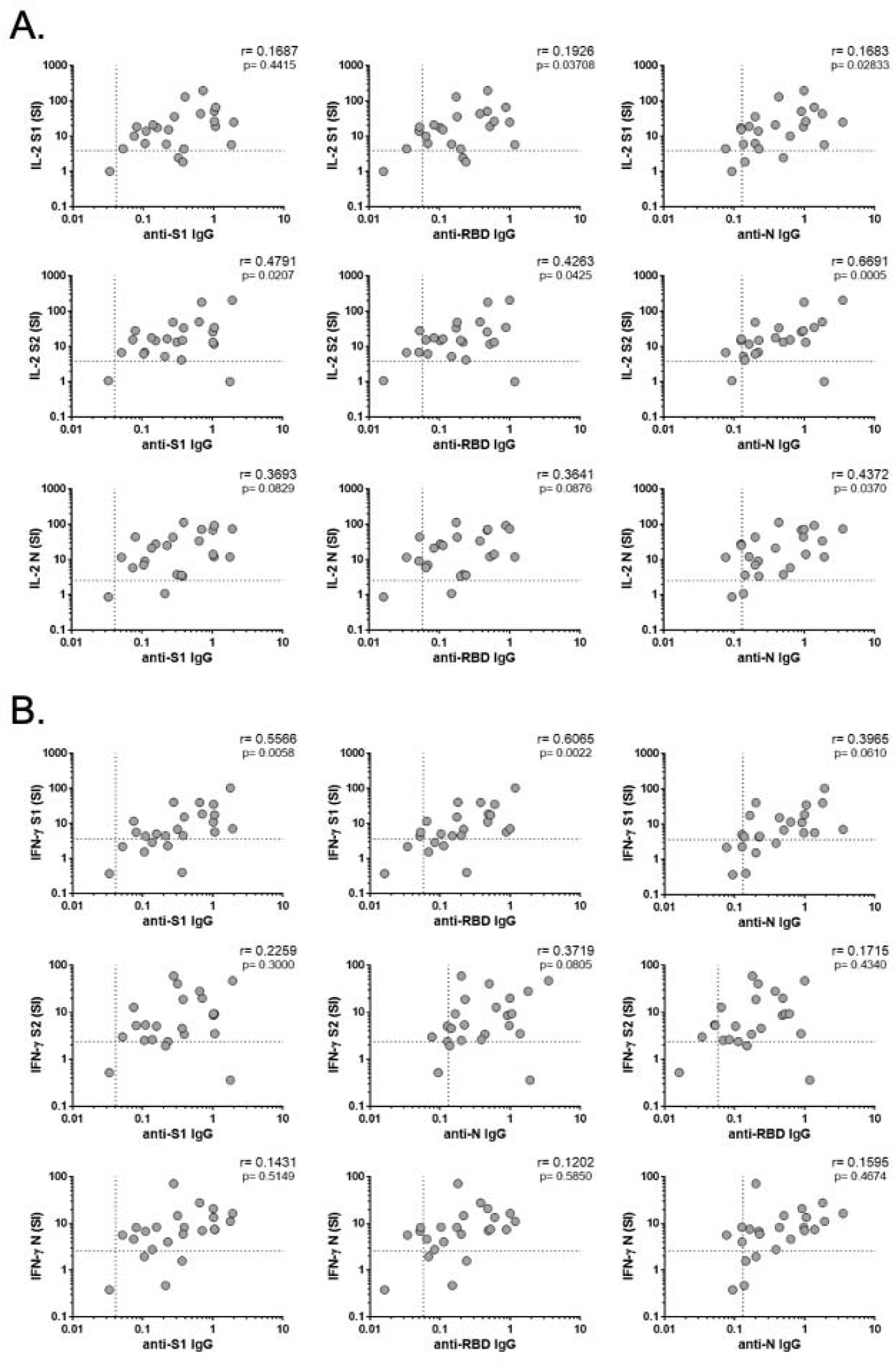
Correlations of IL-2 and IFN-γ stimulation index (SI) with SARS-CoV-2 specific IgG. Linear regressions of (A) IL-2 or (B) IFN-γ stimulation index (SI) in whole blood stimulation assays from COVID-19 recovered individuals (n=23) in response to PepMix S1, S2, and N peptide pools, and SARS-CoV-2 specific IgG (OD values) for S1, RBD and N. Dashed lines represent the cut-off values (IL-2 and IFN-g calculated by ROC analysis; ELISA IgG 2SD obtained from the pre-pandemic population). The r values were calculated using Pearson’s Correlation Test.

### Supplemental Tables

**Supplemental Table I.**
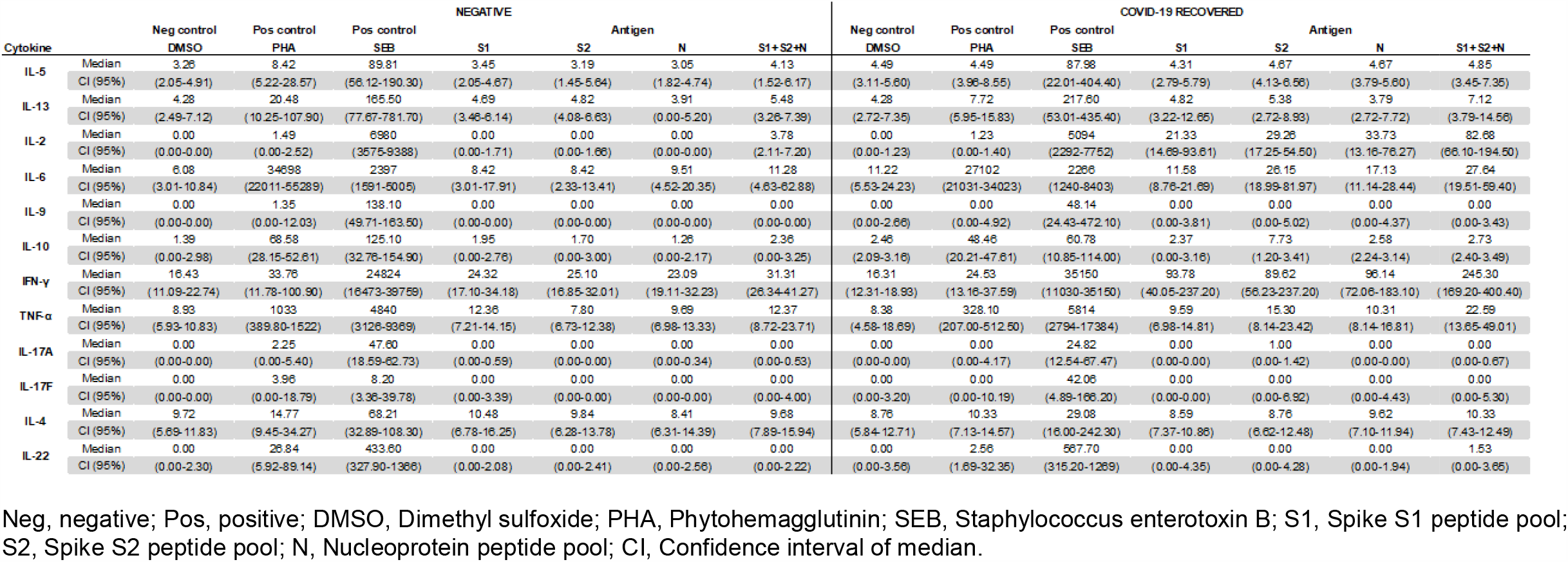
Cytokine production quantified in pg/mL after WBS assays in negative individuals and COVID-19 recovered patients in response to negative (DMSO) and positive controls (10 μg/mL PHA; 1 μg /mL SEB) and to SARS-CoV-2 PepMix peptide pools (1 μg/mL per peptide).

**Supplemental Table II.** ROC curves, cut-offs, sensitivity, and specificity for IL-2 and IFN-γ in the determination of negative and COVID-19 recovered individuals after a WBS assay

| Cytokine |  | Antigen |  |  |  |
| --- | --- | --- | --- | --- | --- |
|  |  | S1 | S2 | N | S1+S2+N |
| IL-2 | Area | 0.9474 | 0.9634 | 0.9497 | 0.9634 |
|  | p value | < 0.0001 | < 0.0001 | < 0.0001 | < 0.0001 |
|  | cut-off | > 3.895 | > 3.760 | > 2.525 | > 9.145 |
|  | Sensitivity (%) (95% CI) | 87 (20/23) | 91.3 (21/23) | 91.3 (21/23) | 91.3 (21/23) |
|  | Specificity (%) (95% CI) | 94.7 (18/19) | 100 (19/19) | 100 (19/19) | 100 (19/19) |
| IFN-γ | Area | 0.8490 | 0.8307 | 0.8741 | 0.9050 |
|  | p value | 0.00012 | 0.00026 | < 0.0001 | < 0.0001 |
|  | cut-off | > 3.630 | > 2.370 | > 2.575 | > 7.730 |
|  | Sensitivity (%) (95% CI) | 73.9 (17/23) | 87 (20/23) | 82.6 (19/23) | 82.6 (19/23) |
|  | Specificity (%) (95% CI) | 94.7 (18/19) | 73.7 (14/19) | 89.5 (17/19) | 94.7 (18/19) |

S1, Spike S1 peptide pool; S2, Spike S2 peptide pool; N, Nucleocapsid; SD, standard deviation

